# Summary of best evidence for interventions for nurse burnout

**DOI:** 10.1101/2024.06.27.24309626

**Authors:** Liping Wang, Guixiang Li, Jiayi Liu, Yanjun Diao, Yu Zhuo

**Affiliations:** West China Hospital, Sichuan University/West China School of Nursing, Sichuan University a Pancreatitis Center; Department of Integrated Traditional Chinese and Western Medicine; Mental Health Center, Sichuan Chengdu 610041

**Keywords:** Nurse, Job burnout, Intervention, Evidence-based nursing, Evidence summary

## Abstract

**Objective:** To search, evaluate and summarize the best evidence on nurse burnout intervention at home and abroad, and provide evidence-based basis for clinical nursing staff.

**Methods:** The literature on nurse burnout intervention was searched in Chinese and English guideline websites and databases, including guidelines, evidence summaries, systematic reviews, clinical decision-making, expert consensus, and randomized controlled studies. The search period was from database establishment to December 31, 2023.

**Results:** 16 articles were included, including 1 guideline, 1 consensus, 1 evidence summary, 1 randomized controlled study, and 12 systematic reviews. After comprehensive analysis and induction, we extracted five aspects, including effective communication, self-care, resource support, education and training, and social environment, with a total of 15 best evidence.

**Conclusion:** The best evidence for evidence-based intervention for nurse burnout is scientific and practical, and can provide a certain reference for clinical practice in reducing nurse burnout.

---

Burnout is usually a response to chronic occupational stressors, characterized by increased emotional exhaustion and depersonalization, and decreased personal accomplishment ^[1]^. Nurses have problems such as high intensity, fast pace, high risk of occupational exposure, and frequent emergencies ^[2]^, which makes them more likely to suffer from burnout than other medical staff ^[3]^. It is reported that the burnout rate of nurses abroad is 40.0% ^[4,5]^, while the burnout rate of nurses in China is 64.5 % ^[6]^, which is much higher than that in Europe and the United States. The COVID-19 pandemic has exacerbated the burnout rate of nurses, and foreign studies have shown that it is as high as 68 % ^[7]^. Burnout is a serious occupational health problem. Nurses with burnout may experience physical and psychological symptoms such as fatigue, anxiety, depression, and overall job dissatisfaction ^[8]^, and are more likely to abuse substances including caffeine, alcohol, and illegal drugs ^[9]^. At the same time, burnout has also been shown to have a negative impact on individuals’ interpersonal relationships and family life, and is associated with a higher incidence of physical diseases such as hypertension, heart disease, and sleep disorders ^[10]^. Nurses with high levels of burnout can cause negative effects such as hospital-acquired infections, medication errors, and patient falls ^[11]^. Therefore, it is increasingly important to actively intervene in nurse burnout. This study uses relevant research on interventions for nurse burnout as evidence, evaluates, summarizes, and provides a reference for clinical nursing workers.

## 1 Materials and methods

### 1.1 Identify the problem

Evidence-based questions were raised according to the PICO principle, where P (population) refers to nurses experiencing burnout; I (intervention) refers to nurse burnout intervention and management; C (comparison) refers to conventional burnout intervention measures; and O (outcome) includes nurses’ job stress, nursing quality, turnover rate, burnout intervention, etc.

### 1.2 Search strategy

According to the "6S" evidence model, the keywords "nurse* /paramedic* /nursing staff ", " job burnout /burnout /emotional exhaustion /depersonalization /diminished personal accomplishment ", " prevention / treatment / precautions/intervene / measures/intervention // countermeasures " were searched in the National Guidelines Group (NGC), the Global Guidelines Network (GIN), the Scottish College Guidelines Network (SIGN), the National Institute for Health and Care Excellence (NICE), the Registered Nurses’ Association of Ontario (RNAO), the JBI Centre for Evidence-Based Healthcare Database, BMJ Best Practice, Web of Science, UpTo Date, PubMed, Embase, Cochrane Library, OVID, and CINAHL; the keywords " nurse, nursing staff ", " occupational burnout, work burnout, occupational exhaustion, emotional exhaustion, depersonalization, low sense of accomplishment ", and " intervention, treatment, measures, countermeasures, management" were used to search the commonly used domestic databases and guidelines, such as Biomedicine, CNKI, VIP, Wanfang, and Yimaitong. The retrieval time is from database establishment to December 31, 2023.

### 1.3 Literature inclusion and exclusion criteria

Inclusion criteria: (1) The research subjects were clinical nurses; (2) The research content involved intervention or management of nurse burnout; (3) The type of literature was guidelines, evidence summary, systematic review, expert consensus, randomized controlled trial; (4) The language was English or Chinese. Exclusion criteria: (1) The full text could not be obtained; (2) The literature was included repeatedly.

### 1.4 Literature quality evaluation

(1) Guidelines were evaluated using the AGREE II evaluation tool ^[12]^. (2) Evidence summaries were evaluated using the CASE evaluation tool ^[13]^. (3) Expert consensus, systematic reviews, and randomized controlled trials were evaluated using the Australian JBI Centre for Evidence-Based Healthcare Systematic Review Criteria (2016) ^[14]^.

### 1.5 Screening and integration of evidence

The 2014 version of the JBI evidence pre-grading system ^[15]^ was used to pre-grade the evidence, with the evidence graded from 1 to 5 from high to low. Two researchers with a background in evidence-based nursing independently evaluated the quality of the literature and determined the recommendation level based on the feasibility, appropriateness, and effectiveness of the evidence, namely, level A (strong recommendation) and level B (weak recommendation). If there was a disagreement, a third evidence-based nursing expert intervened to make a judgment to ensure the accuracy of the literature quality evaluation.

## 2. Results

### 2.1 General characteristics of included literature

A total of 637 articles were initially identified, of which 477 were removed after duplicates. After reading the title, abstract, keywords, and full text, 16 articles were finally included ^[16-31]^, including 1 guideline, 1 consensus, 1 evidence summary, 1 randomized controlled trial, and 12 systematic reviews (see Table 1).

**Table 1.**
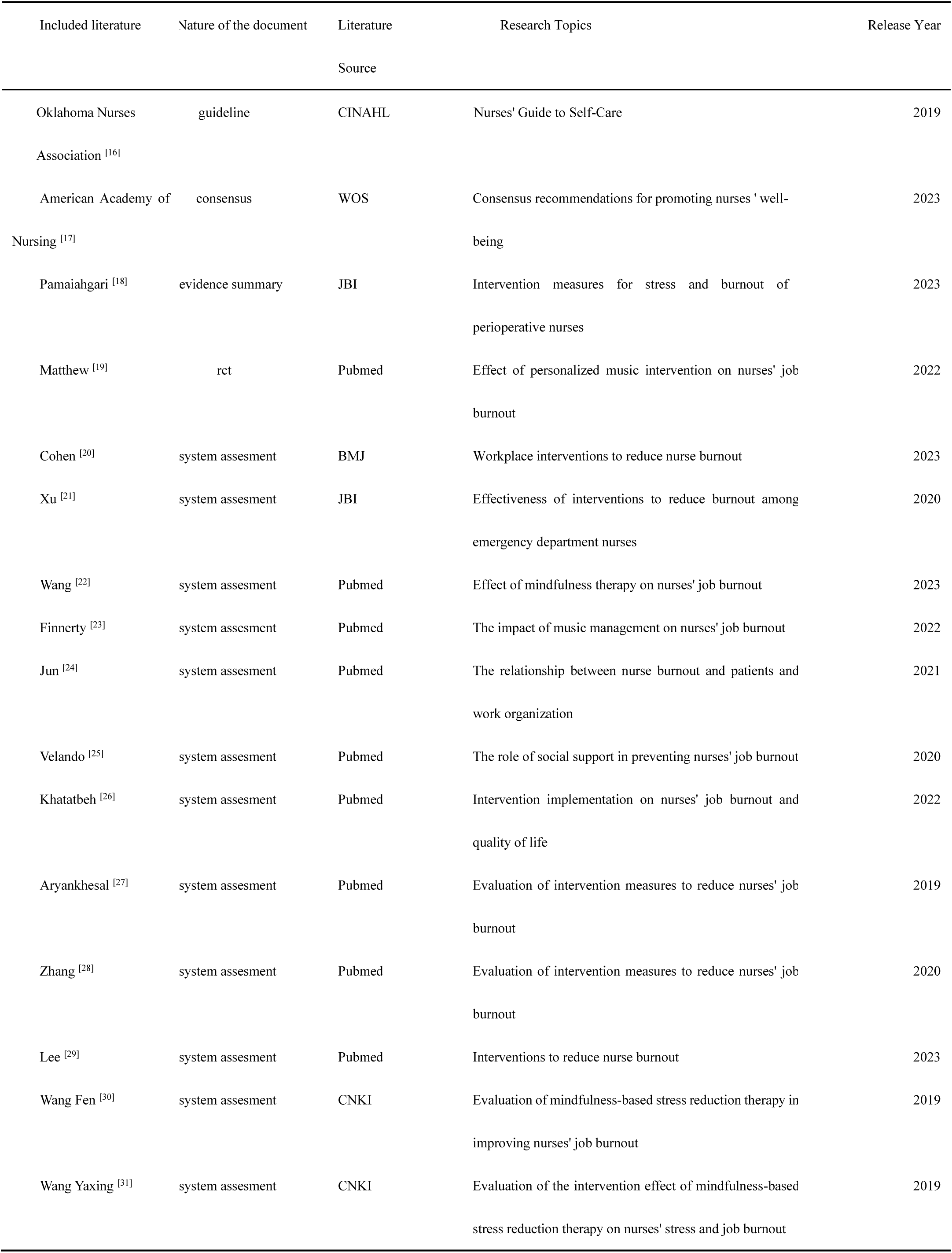
Basic characteristics of included literature (n = 16)

### 2.2 Quality evaluation of included literature

(1) Guidelines: This study included 1 guideline ^[16]^, with 6 areas with a recommendation level of A. The quality of the guideline is high and recommended for use. (2) Consensus: This study included 1 consensus ^[17]^, with all items rated “yes” and recommended for use. (3) Evidence summary: This study included 1 evidence summary ^[18]^, with item 3 “ Is the review clear and transparent?” and item 7 “Are the recommendations timely?” rated “no”, and the quality evaluation results of the other items were all “yes” and recommended for use. (4) Randomized controlled trials: This study included 1 randomized controlled trial ^[19]^, with item 5 “Is the interventionist blinded?” and item 6 “Is the outcome assessor blinded?” rated “no”, and the quality evaluation results of the other items were all “yes” and recommended for use. (5) Systematic reviews: This study included 12 systematic reviews ^[20-31]^, with the quality evaluation results shown in Table 2, and all of them were included.

**Table 2.**
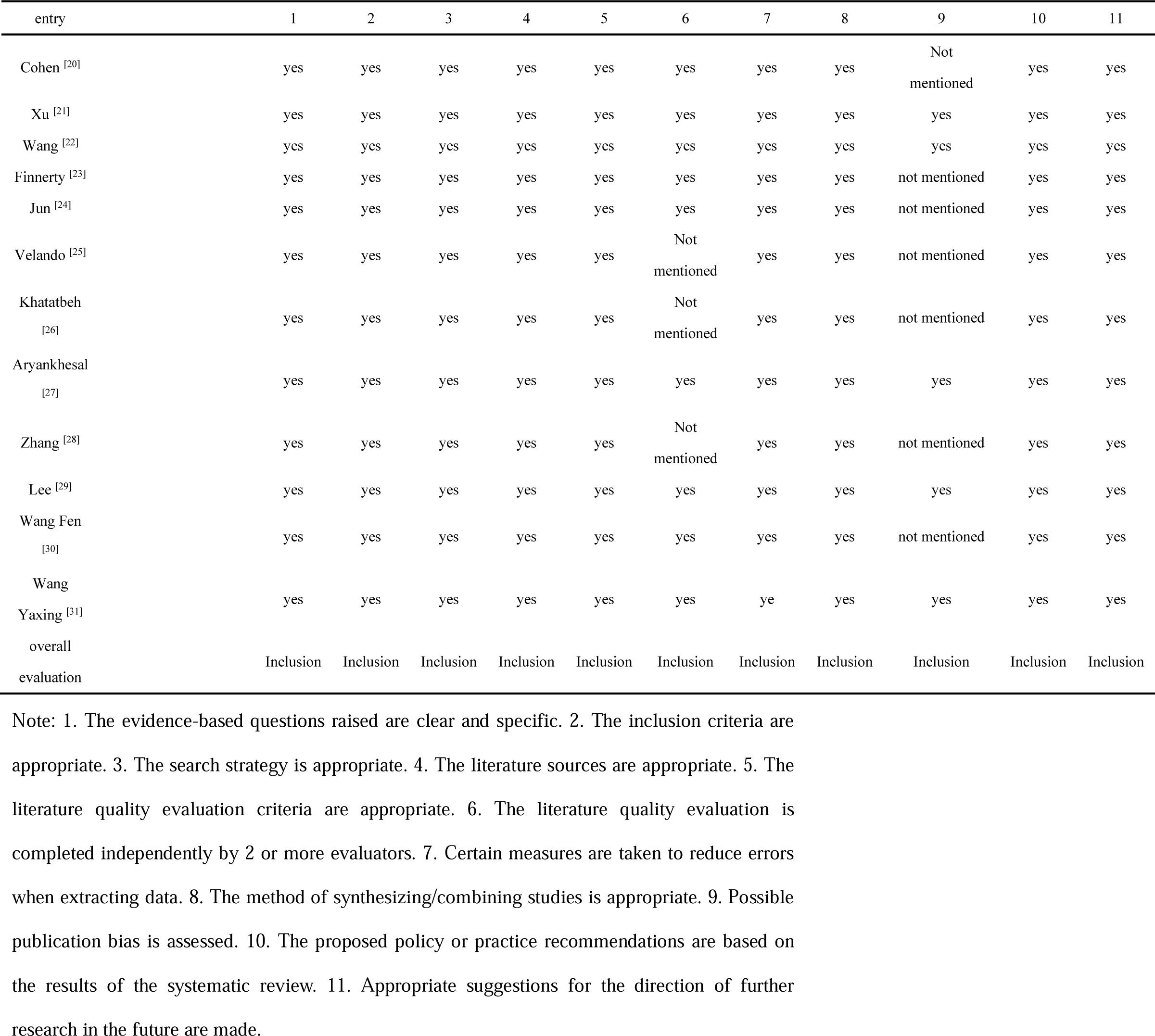
Quality results of included systematic reviews (n = 12)

### 2.3 Evidence summary

This study summarized the relevant literature on nurse burnout intervention, mainly including five aspects: effective communication, self-care, resource support, education and training, and social environment. A total of 15 best evidences are included. The results are shown in Table 3.

**Table 3.** Summary of best evidence for interventions for nurse burnout.

| Evidence topic | Evidence content | Level of evidence | Recommended level |
| --- | --- | --- | --- |
| Effective Communication | 1. It is recommended to strengthen communication among colleagues , analyze and discuss occupational burnout , find out its causes and find solutions . At the same time , training courses can be provided for nurses to improve effective communication skills, help nurses improve their relationships with colleagues and patients, reduce conflicts and stress, and improve work efficiency and satisfaction <sup>[21,27]</sup> | 2 | A |
|  | 2. Encourage nurses to actively participate in work decision-making and problem-solving, actively share their personal ideas and suggestions with managers, enhance nurses' work engagement, and improve their professional identity and sense of belonging <sup>[21]</sup> | 2 | B |
| Self-Care | 3. It is recommended to learn some mental and physical relaxation methods. You can choose mindfulness stress reduction, meditation, self-hypnosis, listening to music and other methods to relax your mind; you can also choose yoga, acupuncture, massage, aerobics and other methods to relax your body. Combining these two methods can help nurses better cope with stress and reduce job burnout <sup>[18-23,27,28,30,31]</sup> | 1 | A |
|  | 4. By writing a gratitude diary or participating in gratitude activities, nurses can transform their negative thoughts into a more positive work attitude, thereby effectively alleviating symptoms of occupational burnout and improving job satisfaction and happiness [20,27]. | 2 | A |
|  | 5. It is recommended to carry out appropriate physical exercise, diet adjustment, sleep management, etc. These measures can improve the physical health of nurses, reduce stress and fatigue, and reduce occupational burnout. [18] | 5 | A |
| Resource Support | 6. Provide nurses with opportunities for continuing education and career development, such as training, further studies, and promotion, to enhance their professional competence and career satisfaction [17] | 5 | B |
|  | 7. Provide psychological support and counseling. Establish a professional burnout counseling team to provide nurses with psychological counseling services to help them relieve work pressure, deal with emotional problems, and enhance psychological resilience. At the same time, positive encouragement and affirmation from leaders and colleagues can<br>effectively improve nurses' sense of efficacy and reduce the occurrence of burnout . [27] | 3 | A |
|  | 8. Actively organize team activities, such as communication meetings , group stress management meetings , etc., to establish a social support network , promote connections between colleagues , and enhance work enjoyment and employee engagement [20,24-26] | 1 | A |
|  | 9. Create a healthy working environment, actively adjust work tasks , motivate management, improve the nurse-patient ratio , optimize work processes, provide flexible work schedules , ensure that nurses have reasonable and adequate working hours, and avoid long, high-intensity work schedules. Provide nurses with adequate vacation and rest time so that they can recover their energy and maintain a good working state. At the same time, training can be used to improve nurses' work efficiency and teamwork ability and reduce their workload [17,21,26,27,29] | 1 | A |
|  | 10. Establishing policies such as nurse family support and assistance programs or parental leave can effectively reduce the family burden of nursing staff, reduce family conflicts, and reduce job burnout. <sup>[16]</sup> | 5 | B |
| Education and Training | 11. Organize individual-themed training , such as self-care workshops, stress management training programs, emotional intelligence education , positive psychology training , cognitive theory training, and career planning, to help nurses better understand and cope with burnout <sup>[16,17,21,28]</sup> | 1 | A |
|  | 12. Organize group -themed training, such as team psychological education training , group face-to-face communication, multidisciplinary seminars, Balint group training, etc., to improve nurses' interpersonal relationship management skills and promote team harmony. <sup>[29]</sup> | 2 | A |
| Social Environment | 13. Strengthen social support and recognition for the nursing profession. Through the media, the Internet and other channels, we should widely publicize the importance of the nursing profession, improve the public's awareness and understanding of the work of nurses, abandon the prejudice that doctors are more important than nurses , optimize public opinion, advocate new professional values, give nurses fair social treatment, respect the nursing profession, and create a good social and public opinion environment for their profession. <sup>[17]</sup> | 5 | B |
|  | 14. Relevant organizations should formulate targeted laws and regulations to clarify the definition of workplace violence in hospitals and impose heavy penalties on nursing staff who commit workplace violence, so as to ensure the occupational safety of nurses. <sup>[16]</sup> | 5 | B |
|  | 15. It is recommended that according to China's actual situation, establishing a reasonable nurse-patient ratio through legislative means can effectively reduce nurses' work pressure, thereby alleviating their job burnout and improving nursing quality. <sup>[17]</sup> | 5 | B |

## 3. Evidence Analysis

### 3.1 Effective Communication

Evidence 1 and 2 clearly affirm the importance of nurses ‘communication skills. Insufficient communication skills may not only lead to increased stress and burnout, but also confusion and loss of confidence among team members ^[32,33]^. Good communication skills are an important skill required in nursing work ^[34]^. Studies have found ^[35]^ that communication skills training has a positive impact on nurse burnout, can improve nurses’ self-efficacy and communication skills in critical situations, directly affect nurses’ mental health, and improve their adaptability and work efficiency, constructively deal with emotional exhaustion and depersonalization, and enhance personal achievement. Surveys shows that compared with hospitals with moderate participation in decision-making, the burnout rate of nurses in hospitals with high participation is 36% lower ^[36]^. Nurses in many countries are dissatisfied with the opportunity to participate in decision-making and complain that leaders do not listen to and respond to their demands ^[37]^. Encouraging nurses to participate in decision-making can help establish a supportive working atmosphere and promote nurses’ autonomous motivation ^[38]^. Leaders or hospital organizations can create an open communication environment, conduct face-to-face communication, and involve nurses in improvement plans to enhance their sense of involvement and identification with their work and reduce the occurrence of burnout.

### 3.2 Self-Care

Evidence 3 to 5 describe the effective role of self-care in coping with burnout. Self-care is generally defined as "engaging in behaviors that maintain and promote physical and mental health, which may include factors such as sleep, exercise, emotion regulation strategies, and mindfulness practices" ^[39]^. The prevention and reduction of burnout should be centered on self-care ^[40]^. The core of mindfulness is self-regulation of attention, focusing on the present moment with curiosity, acceptance, and openness ^[41-43]^. Several studies have found that after nurses took mindfulness courses, their total burnout scores decreased by 12 to 30% ^[44-46]^. Brain imaging studies have also shown that mindfulness training can enhance brain activity, which is related to positive emotions and emotion regulation ^[47,48]^. Writing a gratitude diary or holding a gratitude-themed meeting is also a mindfulness practice that helps to cope with stress and uncertainty at work ^[49]^. It can refocus positive thoughts and energy, bring individuals back to the source of their happiness, allow them to discover the positive factors around them, and reduce burnout ^[20,40]^. Research has found that music intervention has a certain effect on improving nurses’ job burnout^[23]^. Electroencephalograms further showed that both slow-tempo and normal-tempo music reduced rapid brain activity in the right frontal lobe, suggesting that music can be an effective means of reducing negative emotions ^[50]^. It is recommended that nurses choose appropriate ways to manage stress and job burnout in their daily lives and medical environments to promote their own health.

### 3.3 Resource Support

Evidence 6 to 10 describe the necessity of providing nurses with certain resource support at work. Social support is considered a work resource that can be used to cope with work demands and prevent the occurrence of burnout ^[51]^. Social support can come from family or friends to help nurses reduce the impact of work-family conflict ^[52]^, or from colleagues and leaders to give them more work affirmation and help. Such support can reduce or avoid negative emotions ^[53]^, reduce the impact of stress on nurses’ professional commitment, and ultimately alleviate burnout ^[54]^. A good working environment means having appropriate autonomy, sufficient staff and resources, and good working relationships with doctors and managers. When these conditions are met, the chance of nurse burnout will be reduced by 28% ^[55]^. Studies have found that in hospitals with poor staffing conditions, the possibility of nurse burnout is 78% higher than that in hospitals with better staffing conditions ^[56]^, while another study found that in an environment with adequate staff and support services, a reasonable workload, and sufficient 30-minute rest time, the level of nurse burnout is significantly lower ^[57]^. The concept of "magnetic hospital " has been implemented and certified abroad, and its role in nursing has been recognized. It has many characteristics of a good working environment, such as increased nurse autonomy, better staffing, and supportive management ^[58,59]^. It is recommended that domestic hospitals carry out the construction of magnet hospitals in order to reduce the occurrence of nurse burnout.

### 3.4 Education and Training

Evidence 11 to 12 introduces that individual and group-based education and training can help reduce nurses’ occupational burnout. Group psychological education and training is a training method that focuses on humanistic care. It aims to promote communication and positive interaction among nursing staff, so that nurses can examine problems from different perspectives and adjust their cognition of adverse events from a personal level. Such training helps to improve nurses’ adaptability to the environment and stress resistance, and can effectively reduce occupational burnout ^[60]^. At the same time, a domestic study also showed ^[61]^ that group psychological education and training, as an effective nursing management method, not only helps to improve nurses’ psychological resilience, but also significantly reduces their occupational burnout, so it is worth implementing widely. Emotional intelligence is an individual-based education and training, which refers to the ability of individuals to recognize, express, understand, regulate and manage their own and others’ emotions through scientific training ^[62]^. It is usually manifested as good social adaptability and emotional control, and has a positive effect on reducing nurses’ occupational burnout and stabilizing the nursing team ^[63,64]^. A study conducted a phased emotional intervention on 36 nurses in the emergency department. After comparison, it was found that there were significant differences in the nurses’ mental health, job burnout, and stress coping methods before and after the intervention ^[65]^, indicating that emotional therapy can effectively improve the mental health of nurses ^[66]^. Therefore, appropriate individual and group-based education and training can enable nurses to better understand and deal with job burnout, adjust their stress coping methods, and reduce its adverse effects.

### 3.5 Social Environment

Evidence 13 to 15 illustrate the importance of social environment in nurse burnout. Some countries have proposed coping strategies for nurse burnout from a legal perspective and explicitly require the establishment of a reasonable nurse-patient ratio. For example, Victoria (Australia) and California (USA) have implemented a "minimum nurse-patient ratio" system. Depending on the department, the nurse-patient ratio is usually set between 1: 2 and 1: 4 ^[67]^. Research results show that establishing a reasonable nurse-patient ratio through legislation can effectively reduce the degree of nurse burnout and further improve the quality of nursing work ^[68]^. At present, the concept of valuing doctors over nurses is relatively common in society. The social and economic status of nurses is relatively low, which has seriously undermined their work enthusiasm, leading to an increase in nurse burnout and turnover. Some developed countries have introduced relevant policies and regulations to protect the legitimate rights and interests and physical and mental health of nurses ^[69]^. However, China’s relevant beneficial policies are often difficult to implement due to differences in local conditions. Therefore, it is hoped that more health administrative agencies and associations will pay attention to nurses’ occupational burnout, establish practical and effective management mechanisms to support the nursing work of various units, recognize and positively evaluate their work, enhance positive social public opinion, and create a good social environment for nurses.

## 4 Conclusion

This study summarizes the best evidence of nurse burnout and recommends that clinical nursing staff and managers choose appropriate intervention methods from five aspects: effective communication, self-care, resource support, education and training, and social environment, in order to reduce the adverse effects of nurse burnout. Since the literature included in this study is mainly in English, there are few literature studies on nurse burnout intervention in China. In the future, we will combine relevant actual research in China to conduct an in-depth discussion on the evidence effect of nurse burnout intervention.

The author declares no actual or potential conflicts of interest

## Data Availability

All data produced in the present study are available upon reasonable request to the authors

https://www.example.com

